# Signatures of Cysteine Oxidation on Muscle Structural and Contractile Proteins Are Associated with Physical Performance and Muscle Function in Older Adults: Study of Muscle, Mobility and Aging (SOMMA)

**DOI:** 10.1101/2023.11.07.23298224

**Authors:** Nicholas J. Day, Shane S. Kelly, Li-Yung Lui, Tyler A. Mansfield, Matthew J. Gaffrey, Jesse B. Trejo, Tyler J. Sagendorf, Kwame Attah, Ronald J. Moore, Collin M. Douglas, Anne B. Newman, Stephen B. Kritchevsky, Philip A. Kramer, David J. Marcinek, Paul M. Coen, Bret H. Goodpaster, Russell T. Hepple, Peggy M. Cawthon, Vladislav A. Petyuk, Karyn A. Esser, Wei-Jun Qian, Steven R. Cummings

## Abstract

Oxidative stress is considered a contributor to declining muscle function and mobility during aging; however, the underlying molecular mechanisms remain poorly described. We hypothesized that greater levels of cysteine (Cys) oxidation on muscle proteins are associated with decreased measures of mobility. Herein, we applied a novel redox proteomics approach to measure reversible protein Cys oxidation in vastus lateralis muscle biopsies collected from 56 subjects in the Study of Muscle, Mobility and Aging (SOMMA), a community-based cohort study of individuals aged 70 years and older. We tested whether levels of Cys oxidation on key muscle proteins involved in muscle structure and contraction were associated with muscle function (leg power and strength), walking speed, and fitness (VO_2_ peak on cardiopulmonary exercise testing) using linear regression models adjusted for age, sex, and body weight. Higher oxidation levels of select nebulin Cys sites were associated with lower VO_2_ peak, while greater oxidation of myomesin-1, myomesin-2, and nebulin Cys sites was associated with slower walking speed. Higher oxidation of Cys sites in key proteins such as myomesin-2, alpha-actinin-2, and skeletal muscle alpha-actin were associated with lower leg power and strength. We also observed an unexpected correlation (r = 0.48) between a higher oxidation level of 8 Cys sites in alpha-actinin-3 and stronger leg power. Despite this observation, the results generally support the hypothesis that Cys oxidation of muscle proteins impair muscle power and strength, walking speed, and cardiopulmonary fitness with aging.

## 1. Introduction

Aging is associated with diminished muscle and physical function which leads to mobility disability, loss of independence, and poorer quality of life (de Vries et al., 2012). However, the underlying molecular changes contributing to a decline in muscle function and physical performance remain unclear. In muscle, mitochondria and enzymes like NADPH Oxidases (NOX) generate reactive oxygen species (ROS) such as superoxide radical (O_2_ ^•−^) and hydrogen peroxide (H_2_O_2_) (Bouviere et al., 2021). Studies have shown that ROS are important signaling molecules for underlying muscle function, including modulation of glucose transport (Henríquez-Olguin et al., 2019), insulin sensitivity (Xirouchaki et al., 2021), and mitochondrial function (Kramer, Duan, Qian, & Marcinek, 2015). Indeed, oxidants produced during exercise are essential for the exercise adaptive response, improving antioxidant capacity, and mitochondrial biogenesis (Powers et al., 2020). However, excessive or chronic exposure to ROS associated with aging causes fatigue, reduced contractile function of muscles, and blunted adaptive signaling (Andrade, Reid, Allen, & Westerblad, 1998; Musci, Hamilton, & Linden, 2019).

Excess production of ROS and diminished antioxidant capacity leads to a redox imbalance, also known as “oxidative stress”, which contributes to the etiology of many age associated pathologies (Sies, 2020). However, research in recent decades has shown that the concept of ROS-induced “oxidative damage” is a limited view of ROS and aging (Gladyshev, 2014). The original concept of ROS-based “damage” is really a potentially reversible “oxidative modification” that alters mechanisms of physiological response and functional regulation, rather than causing dysfunction and degeneration. Consequently, the “redox stress” hypothesis has emerged as a theory to explain the relationship between ROS and aging and posits that a shift to a more pro-oxidizing redox state is a causal effect of the aging process (Sohal & Orr, 2012). At the molecular level, ROS-mediated redox signaling can occur through reversible post-translational modifications (PTMs) of the thiol group on protein cysteine residues (Brandes, Schmitt, & Jakob, 2009). These redox PTMs modulate the activity, function, localization, and binding interactions of proteins, and their reversibility enables “switch”-like behavior for transient signaling roles (Brandes et al., 2009). Reversible PTMs on protein cysteine thiols have been increasingly considered as a major regulatory mechanism in oxidative stress-related diseases and aging (Kehm, Baldensperger, Raupbach, & Höhn, 2021).

Mitochondrial dysfunction has been considered a hallmark of aging (López-Otín, Blasco, Partridge, Serrano, & Kroemer, 2023). This situation enhances the production of ROS in aging tissues, resulting in a propensity towards an increasingly pro-oxidizing redox state, leading to “redox stress”. The duality of ROS as both essential, but potentially detrimental for biological functions makes it of particular interest for studying the intersection of aging and muscle function, however identification of proteins that are modified by ROS in human aging muscle has not been investigated. The sarcomere is the fundamental contractile unit in muscle that is responsible for generating force. Research over the last 30 years has demonstrated that the force capacity of the sarcomere is an emergent property of all the proteins that contribute to the structure (Lehman, Crocini, & Leinwand, 2022). Consequently, changes or mutations in any one protein are sufficient to impact the function of the structure, such as those comprising the thick and thin filaments or the Z- and M-lines.

We hypothesize that the levels of cysteine oxidation on sarcomere proteins involved in muscle contraction and structure are associated with physical performance (walking speed), muscle function (leg strength and leg power), and cardiopulmonary fitness (VO_2_ peak). Previous studies have shown that cysteine oxidation of sarcomere proteins can impact the function of the contractile unit. One example is based on the opposing effects of glutathionylation and nitrosylation on Cys 134 of fast-twitch troponin I, which modulates its affinity for Ca^2+^ and subsequent force generation (Dutka et al., 2017). The large protein titin is also subject to oxidative modifications that modulate its mechanical properties such as elasticity and stiffness (Giganti, Yan, Badilla, Fernandez, & Alegre-Cebollada, 2018). To test our hypothesis, we applied a novel redox proteomics approach to quantitatively profile the levels of oxidation on protein cysteines in muscle tissue of older adults to identify signatures that may distinguish among varying degrees of physical performance.

## 2. Results

### 2.1 Study cohort

The Study of Muscle, Mobility and Aging (SOMMA) was conceived to investigate how biological changes in muscle contribute to the decline in mobility – strength and walking – with aging. SOMMA recruited participants aged 70 years or older for extensive tests of physical performance (phenotypes) and collection of biospecimens (Cummings et al., 2023)(**Fig. 1**). Needle biopsies from vastus lateralis muscle were collected for a variety of analyses, such as gene expression, mitochondrial respirometry (Mau et al., 2023), and protein cysteine oxidation (this study). Participants in this exploratory study selected from the larger SOMMA cohort are representative of both sexes, age range, and range of phenotypes measured at the clinical sites. Participant characteristics of this study are summarized in **Table 1**, which were used for testing relationships between site-specific cysteine oxidation and phenotype data related to mobility.

**Figure 1.**
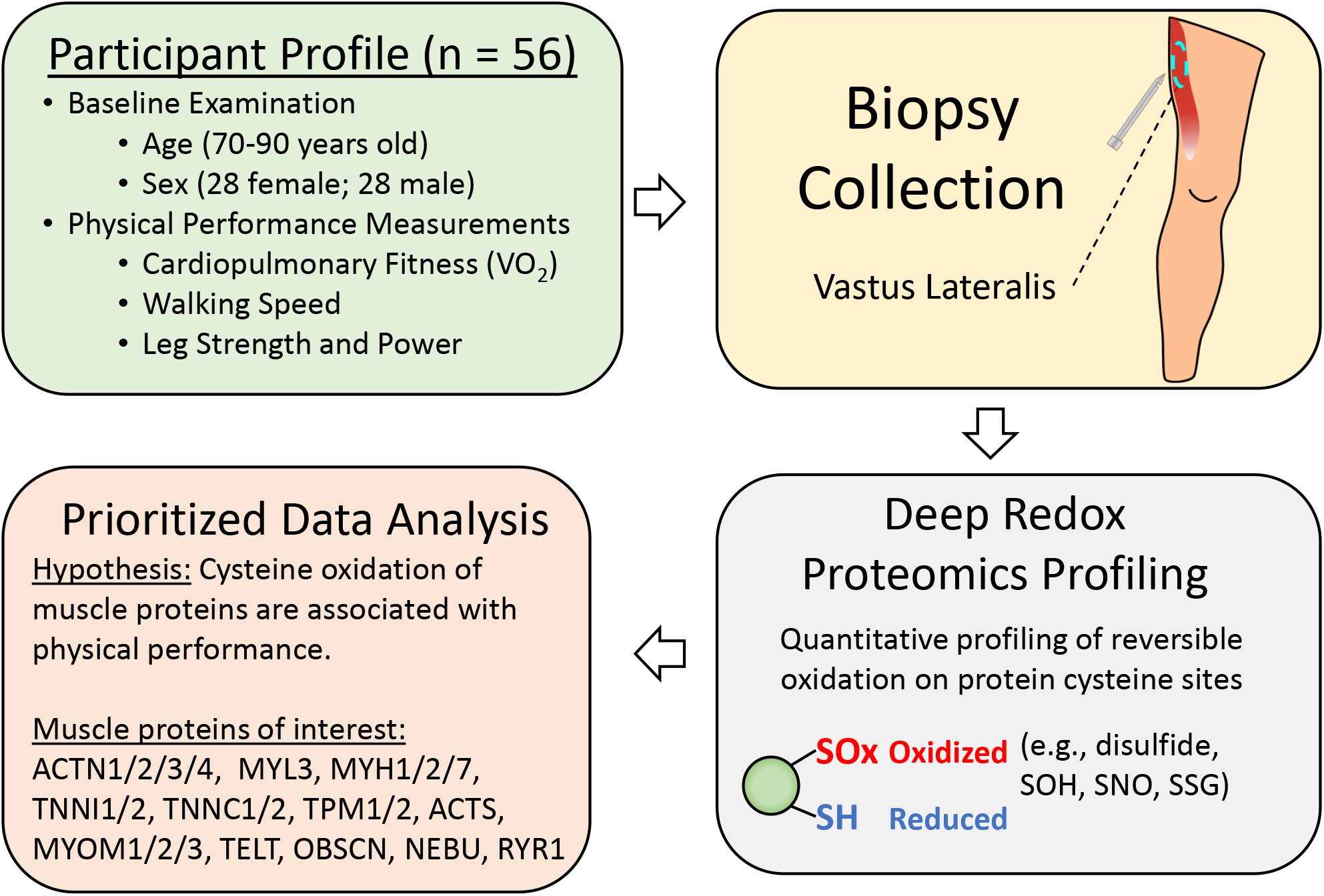
Study design and workflow. Participants undergo baseline physical examinations (age, height, weight, blood pressure, etc.) and measures of muscle function (leg strength and power), walking speed, and fitness (VO_2_ peak) as well as specimen collections (blood, urine, muscle, etc.) over multiple days (Cummings et al., 2023). Muscle biopsies were collected from vastus lateralis muscle. A deep redox proteomics profiling workflow was used to quantitatively measure Cys oxidation (i.e., all forms of reversible thiol PTMs). Analysis of redox proteomics data was prioritized for a select group of muscle contraction proteins (Uniprot ID format) to test the hypothesis that protein Cys oxidation is associated with functional measures.

**Table 1:** Participant characteristics (n = 56)

| Characteristics | Group average |
| --- | --- |
| Age, years | 77.6 ± 5.1 |
| Sex | 28 Men + 28 Women |
| Weight, kg | 79.0 ± 18.8 |
| VO<sub>2</sub> peak, mL/min | 1588.0 ± 498.1 |
| Walking Speed, m/s | 1.0 ± 0.2 |
| Leg Strength (One Repetition Maximum, kg) | 179.3 ± 67.2 |
| Peak leg power, Watts | 347.1 ± 185.8 |

All characteristics reported as mean ± SD

### 2.2 Muscle redox proteome profiling

To test the hypothesis that protein Cys thiol oxidation on specific Cys residues of muscle proteins are associated with reduced muscle function and physical performance, we quantitatively profiled the redox proteome of muscle tissue from older adults. For reference, we define the term “cysteine oxidation” as the total oxidation level of all reversible forms of PTMs on a given Cys residue, which serves as an overall measure of the redox state for a given Cys site (Zhang, Gaffrey, Li, & Qian, 2021). Since the levels of oxidation on protein Cys residues are impacted by factors such as protein abundance, we devised a peptide-level enrichment strategy that is adapted from our previously reported deep redox profiling workflow (Day et al., 2022). This workflow measures both global protein abundance and cysteine oxidation (**Fig. 1; Supplemental Fig. S1A**). To conduct an initial comparison of cysteine oxidation and protein abundance, we performed the first experiment using only 14 tissue samples. Statistical analysis using linear regression models adjusted for sex, age, and weight (see methods) was applied to test associations between protein Cys oxidation and muscle function (leg power and strength), walking speed, and fitness (VO_2_ peak). The results revealed a greater number of significant associations (p < 0.05) for Cys oxidation (122 Cys sites) than protein abundance (15 proteins) (data not shown). These initial experiment results confirm that redox proteome profiling of Cys oxidation provides more informative measurements for testing associations with muscle function than protein abundance. Thus, we focus on quantitative profiling of protein Cys oxidation in muscle tissues from 56 SOMMA participants (**Table 1**).

We used a tandem mass tag (TMT) multiplexed quantification strategy (**Supplemental Fig. S1B**), to allow for multiplexed quantification of Cys oxidation across 14 samples in each multiplex experiment. In total, we measured oxidation of 15,049 Cys sites, covering 4,354 proteins. Using this dataset, we applied a 75% completeness threshold to analyze Cys sites with higher coverage of oxidation data across all plexes, yielding 7,721 Cys sites from 2,657 proteins. Principal component analysis (PCA) highlights the variation in protein oxidation profiles among participants, where most are dispersed throughout the plot with minimal separation due to variables such as sex (**Supplemental Fig. S2**).

### 2.3 Statistical association analysis of Cys oxidation on key muscle proteins

To test our hypothesis, we prioritized the analysis of a select group of 22 muscle structural and contractile proteins that are key components of sarcomeres to determine the association of Cys oxidation on these proteins with four measurements of physical performance (**Fig. 1; Supplemental File S1**). Linear regression LIMMA models (Smyth, 2005) using sex, age, and weight as covariates were used to test associations, where the significance threshold was set at a p value < 0.05 (**Supplemental File S2)**. For each functional measurement, the levels of oxidation at significantly associated Cys sites (p < 0.05) were plotted as heatmaps, which revealed clusters of Cys sites with similar oxidation profiles (e.g., **Supplemental Fig. S3**). We further define each of the clusters as a “signature” of Cys sites. The statistical significance of each signature (or cluster of Cys sites) in association with a given functional measure was further tested by LIMMA, where all signatures were statistically significant (adjusted p < 0.05) (**Supplemental File S3**).

### 2.4 Signatures of Cys oxidation associated with functional measures

We first examined the signatures of Cys sites that show statistically significant associations with VO_2_ peak, which is considered the gold standard measure of cardiorespiratory fitness. A heatmap of all 19 Cys sites that passed our statistical significance cutoff (p value < 0.05) revealed that oxidation levels on specific Cys sites of nebulin (NEBU), obscurin (OBSCN), and myomesin-2 (MYOM2) were associated with cardiopulmonary fitness (**Supplemental Fig. S3**). Pearson correlation was used to further test the significance of the signatures with a given phenotype. The correlation plots of each cluster are shown in **Supplemental Fig. S4**, where clusters 2 and 4 showed a significant negative association (*R* < −0.30; p < 0.05). Among the clusters, one of the NEBU signatures that consisted of 5 Cys sites showed a predominantly negative association with VO_2_ peak (**Fig. 2A and Fig. 3A)**, suggesting the oxidation levels on specific Cys sites of NEBU are functionally related to overall cardiopulmonary fitness.

**Figure 2.**
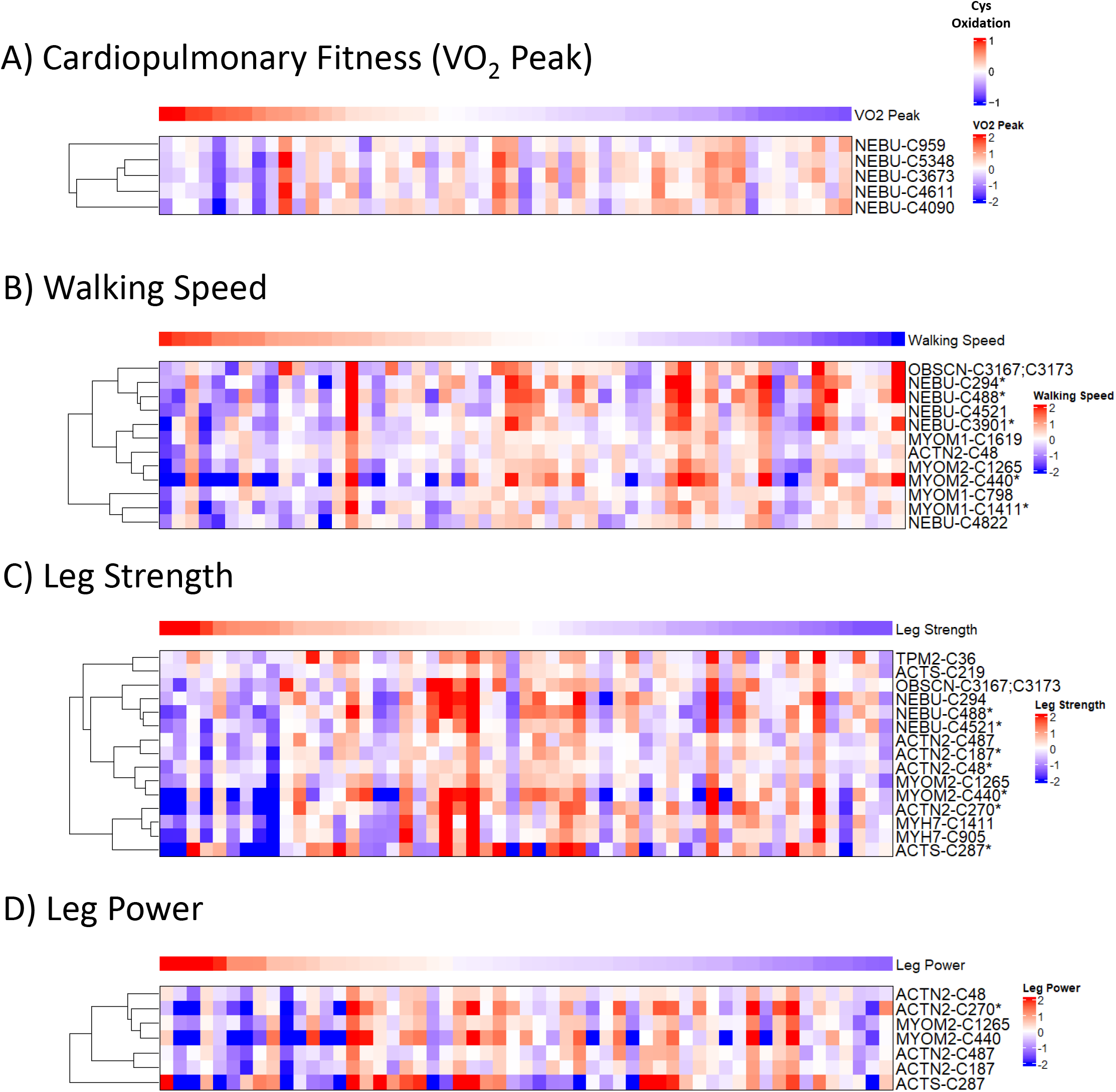
Selected signatures of Cys oxidation in significant association with fitness, muscle function, and physical performance. Cys sites on muscle proteins with oxidation levels significantly associated with a functional measure (**Supplemental File S2**) were clustered and plotted in heatmaps (**Supplemental Figures S3, S5, S7, S9**). A representative cluster of Cys sites (“signature”) from each phenotype-specific heatmap are shown in panels A-D. Each row represents a Cys site with oxidation levels that are significantly associated with a phenotype, while each column represents a participant. Columns are ranked in descending order from left to right based on phenotypic measurement and the phenotypic values are represented by median-centered Z-scores, while Cys oxidation levels were scaled by median-centering. Note that the row-wise hierarchical clustering of Cys sites is reordered from the original corresponding clusters in **Supplemental Figures S3, S5, S7, S9**. Protein identities are in UniProt format. Sites passing an adjusted p value < 0.05 cutoff are denoted by ‘*’. A) Peak volume of oxygen consumption (VO_2_ peak; mL/min) as a measure of cardiopulmonary fitness. B) Walking speed from a 400m walking test (m/s). C) Leg strength as measured by one repetition maximum (kg). D) Leg power measured as the highest peak power (watts) generated.

**Figure 3.**
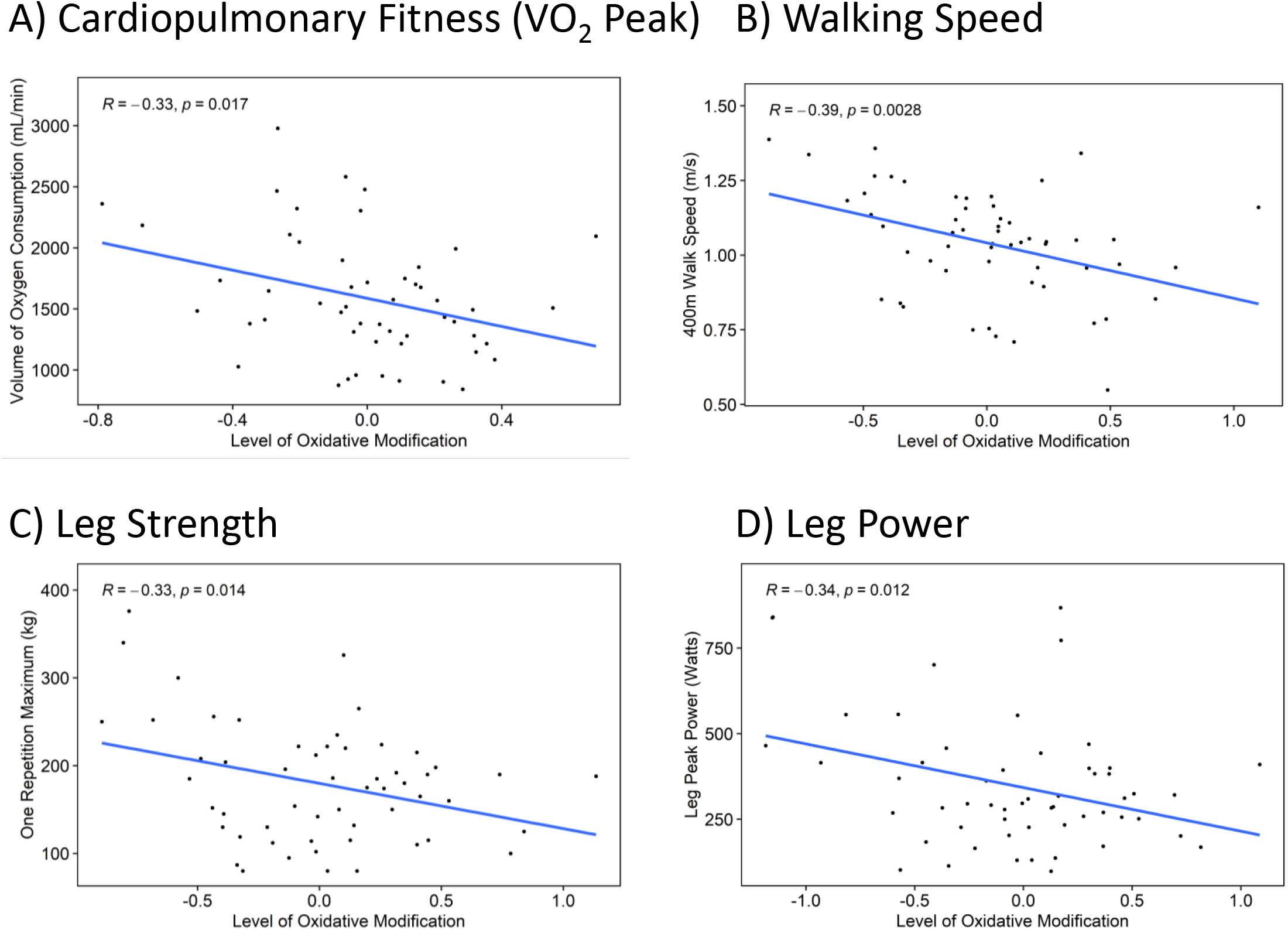
Selected signatures of Cys oxidation are negatively associated with fitness, muscle function, and physical performance in older adults. Signatures (sub-clusters) within each phenotype-specific heatmap (**Supplemental Figures S3, S5, S7, S9**) were plotted for correlation analysis (**Supplemental Figures S4, S6, S8, S10**). Representative correlations of signatures of highlighted in **Fig. 2** are shown in panels A-D, where the mean level of oxidation across all Cys sites in a signature were plotted for each participant against their phenotypic measurement. *R* and p values are derived from Pearson correlation. A-D: Representative Pearson correlations for A) VO_2_ peak, B) walking speed, C) leg strength, and D) leg power.

Time to complete a 400-meter walk is a key metric of mobility in older adults, thus we next tested associations between protein Cys oxidation and average walking speed. As shown in the overall heatmap (**Supplemental Fig. S5**), a total of 35 Cys sites were associated with walking speed, which include 14 out of the 22 proteins being tested (**Supplemental File S1**). Correlation analysis (**Supplemental Fig. S6**) indicated that all clusters except cluster 5 displayed significant, negative associations with walking speed (*R* < −0.30; p < 0.05). **Fig. 2B** shows a signature of 12 Cys sites (i.e., cluster 2 of **Supplemental Fig. S5**) corresponding to myomesin-1 (MYOM1), alpha-actinin-2 (ACTN2), MYOM2, NEBU, and OBSCN that were significantly associated with walking speed. The negative correlation (*R* = −0.39, p = 0.028; **Fig. 3B**) suggested that there is a negative association between Cys oxidation of these proteins and walking speed.

We next studied the association between leg strength, leg power, and Cys oxidation. As shown in the heatmap (**Supplemental Fig. S7**), a total of 41 Cys sites from 15 proteins displayed different levels of association with leg strength. Correlation analysis of each cluster (**Supplemental Fig. S8**) revealed that only clusters 5 and 6 had significant negative associations (*R* < −0.30; p < 0.05). **Fig. 2C and 3C** highlights the signature of Cys sites (cluster 6 from **Supplemental Fig. S7**; *R* = −0.33, p = 0.014) from several proteins such as the alpha isoform of skeletal muscle actin (ACTS), myosin-7 (MYH7: slow myosin heavy chain), tropomyosin beta chain (TPM2), ACTN2, OBSCN, NEBU, and MYOM2. Furthermore, **Supplemental Fig. S9** shows 33 Cys sites from 11 proteins that were significantly associated with leg power. Clusters 2 and 5 (**Supplemental Fig. S10**) had either a significant negative or positive correlation with peak leg power (*R* < −0.30 or *R* > 0.45; p < 0.05). **Fig. 2D** and **3D** shows a cluster containing 7 Cys sites (cluster 2 of **Supplemental Fig. S9**) corresponding to ACTS, ACTN2, and MYOM2 with a negative association (*R* = −0.34; p = 0.012) between leg power and Cys oxidation.

In agreement with our general hypothesis, most of the clusters exhibited a negative association between Cys oxidation and all functional measures. However, we observed a surprisingly different signature for alpha-actinin-3 (ACTN3), which shows a significant positive correlation (*R* = 0.48, p = 0.00021; **Fig. 4A-B**). This specific signature contains 8 Cys sites that are exclusive to ACTN3, suggesting potential functional regulation of this protein involved in leg power. Interestingly, ACTN2 is a homolog of ACTN3 with roughly 80% identity but does not show a similar relationship between oxidation and leg power (**Fig. 2D**). From a structural standpoint, these sites were mapped to the predicted structure of ACTN3, where we identified 2 pairs of Cys sites within the spectrin repeat region that are potentially in close enough proximity to form structural disulfides (**Fig. 4C**, Cys 490;494 and Cys 593;601). Cys194 is found within the actin binding domain, while Cys788 and 869 are each found within a pair of EF hands that make up the Calmodulin-like domain (**Fig. 4C**), suggesting that redox regulation of Cys residues is a potential mechanism for modulating multiple functional regions of ACTN3.

**Figure 4.**
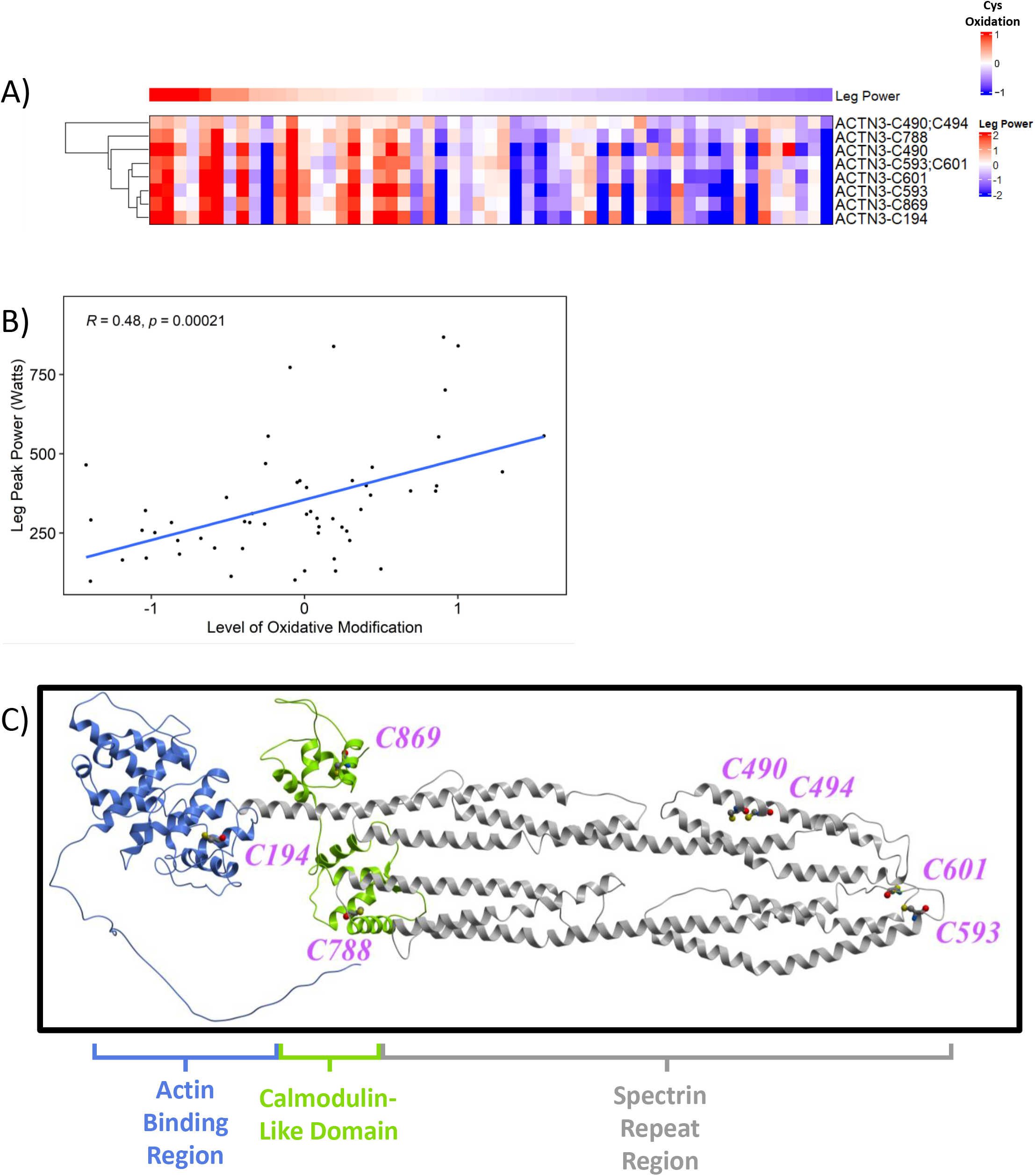
Unique, significant positive association between ACTN3 and leg power. A) Representative heatmap of ACTN3 signatures identified as significantly associated with the leg power phenotype. Each row represents a Cys site with oxidation levels that are significantly associated with a phenotype, while each column represents a participant. Columns are ranked in descending order from left to right based on phenotypic measurement and the phenotypic values are represented by median-centered Z-scores, while Cys oxidation levels were scaled by median-centering. Protein identities are in UniProt format. B) Cys sites from the ACTN3 signature in panel A were plotted for correlation analysis, where the mean level of oxidation across all Cys sites in a signature were plotted for each participant against their phenotypic measurement. *R* and p values are derived from Pearson correlation. C) AlphaFold-predicted structure of ACTN3 with all significantly associated Cys sites denoted by magenta colored text. The actin binding region (AA1-262), neck and spectrin repeat region (AA263-758), and calmodulin-like domain containing 2 pairs of EF hands (AA759-901) are colored in blue, gray, and green, respectively.

## 3. Discussion

Our current study not only provides a first-time view into the molecular landscape of protein Cys oxidation in aging human muscle tissues, but also demonstrates that levels of Cys oxidation on key muscle sarcomeric proteins are generally negatively associated with poorer physical performance. In doing so, the data supports the hypothesis that Cys oxidation signatures are associated with muscle and physical performance. Linear regression and correlation analyses led to identification of signatures with Cys sites whose oxidation levels are significantly associated with muscle function (leg strength and power), physical performance (walking speed), and fitness (VO_2_ peak). These signatures contain several proteins that contribute to sarcomere structure as well as contractile activity such as NEBU, OBSCN, ACTS, MYH7, ACTN2, TPM2, MYOM1, and MYOM2.

For many decades, oxidative stress has been speculated to be a driver of muscle pathologies and aging, however this has not been previously tested in humans. Prior to this study, the relationship between Cys oxidation and aging has been tested in animal models (Campbell et al., 2019; Xiao et al., 2020), but not in humans. The findings presented in this study may begin to unravel the relationship between protein Cys oxidation and muscle function that is emerging as a regulatory mechanism in muscle biology. Given the importance of oxidative stress in the aging process, the application of redox proteomics has value for studying other domains in aging research to explore molecular mechanisms of aging and chronic aging-related diseases.

### 3.1 Implications of Cys oxidation on muscle proteins

Among the structural and contractile proteins that were investigated, oxidation levels of Cys sites on several proteins repeatedly emerged as significantly associated with different measures of muscle function, physical performance, and fitness, emphasizing their potential importance in aging muscle and mobility. One such case is nebulin, where it was found in multiple signatures that were significantly and negatively associated with all four phenotypic measurements (**Supplemental File S2**), along with its exclusive signature for VO_2_ peak (**Fig. 2A**). With a total of 6669 amino acids, nebulin is a giant protein that serves as an integral component of the skeletal muscle thin filament. It contributes to structure through regulation of thin filament length and plays a role in muscle contraction (Slick et al., 2023). Oxidation of Cys sites on this protein may disrupt these essential interactions and dysregulate the organization of actomyosin filaments.

One of the most striking observations in this study revolves around the association of ACTN3 Cys oxidation and leg peak power (**Fig. 4A-B**). Cys868 of ACTN3 has been identified in an aging mouse study as redox-sensitive (McDonagh, Sakellariou, Smith, Brownridge, & Jackson, 2014), which is orthologous to human Cys 869 (**Fig. 4A**), supporting the notion that this protein function could be subjected to redox regulation. Contrary to our hypothesis and the notion that higher Cys oxidation results in a decline of muscle performance, the oxidation levels on multiple Cys sites of ACTN3 are positively correlated with peak leg power (*R* = 0.48). The gene encoding _α_-actinin-3, regarded as the “gene of speed”, is associated with speed and power in athletes, and is exclusively expressed in type-II fast twitch muscle fibers, while ACTN2 is expressed in all skeletal muscle fibers (Pickering & Kiely, 2017). Oxidation in the different functional domains of ACTN3 may serve important roles in regulating its function in type-II fibers responsible for generating maximum muscular power.

In the sarcomere, alpha-actinin proteins bind and crosslink overlapping F-actin filaments at the Z line, making them important proteins for muscle integrity and quality (Burgoyne, Morris, & Luther, 2015). The actinin proteins also serve an important role in the conformational changes of the Z-disc lattice during contraction to propagate force transmission across sarcomeres (Oda & Yanagisawa, 2020). Specifically, the calponin-homology domains of alpha-actinin are important for conformational changes of the protein, where oxidation of Cys194 was observed (Haywood et al., 2016) (actin binding region; **Fig. 4C**). Further, alpha-actinin interacts with titin through its EF-hand domains, where we saw oxidation of Cys788 and Cys869 (Young & Gautel, 2000) (calmodulin-like domain; **Fig. 4C**). Disruption to this interaction has effects on myofilament lattice spacing with subsequent changes in cross-bridge kinetics (Rodriguez Garcia et al., 2023). The difference in oxidation levels at Cys869 within the EF-hand 2 domain of ACTN2 and ACTN3 may partially explain the differences in the negative and positive correlations between power measures and oxidation levels of ACTN2 and ACTN3.

A number of factors could drive the highly contrasting alpha actinin 2 and 3 oxidation profiles, such as the ACTN3 genotype, where the distribution of individuals homozygous for a premature stop codon R577X polymorphism is estimated to be 20% (North et al., 1999). With this frequency, it is possible that some subjects in this study may have limited expression of ACTN3, which would affect the levels of protein Cys oxidation detected in our assay. The ACTN3 genotype may also influence muscle type I slow twitch/ type II fast twitch fiber distribution (Vincent et al., 2007), adding to the list of factors that may explain our results. The exclusive expression of ACTN3 in type II fast twitch muscle fibers also suggests fiber-type specific physiological functions, which have differing responses to oxidants (Anderson & Neufer, 2006) and could play a part in the ACTN3 oxidation profile observed in this study. Subsequent studies on the role of redox-based PTMs on ACTN3 are warranted to gain a better understanding of its functional regulation.

Besides signatures exclusive to single proteins, we also observed instances of oxidized Cys sites on proteins like TNNI2 and ACTS that have been previously reported in the literature. Our assay identified oxidation at Cys134 of fast skeletal muscle troponin I (TNNI2) as significantly associated with other clusters in the walking speed and leg power analyses (**Supplemental File S2**; **Supplemental Fig. S5, S7**). Previous structural analysis work identified this Cys site as solvent-accessible, suggesting it is oxidant-accessible (Gross & Lehman, 2013). Other work revealed that the site undergoes S-Glutathionylation and S-Nitrosylation oxidative modifications (Dutka et al., 2017) that modulate Ca^2+^ sensitivity and force generation, emphasizing this as a redox-sensitive site. Another protein is skeletal muscle alpha actin, which is one of the basic components of the thin filaments in muscle but is also susceptible to Cys oxidation. We identified Cys219 on ACTS as significantly associated with leg strength (**Fig. 2C**), while Cys 287 was significantly associated with both leg strength and leg power (**Fig. 2C-D**). These sites have been identified as redox-sensitive Cys residues in prior work comparing adult and aged mouse skeletal muscle (McDonagh et al., 2014). Oxidation of actin can disrupt interactions with myosin, resulting in attenuated contraction and force production (Elkrief, Cheng, Matusovsky, & Rassier, 2022).

### 3.2 Limitations of this study

This study has several limitations to be acknowledged. While this study focused on measurements of Cys oxidation, global protein abundance profiling could provide additional insight into the biology of impaired muscle performance. Deep profiling of global protein abundances provides more resolution on other aspects, such gene expression or protein degradation, which can help to interpret observations beyond the level of PTMs. With 56 community-based volunteers, the study has limited statistical power to test associations and other confounding factors such as race, metabolism, and lifestyle, which may influence the heterogenous patterns of Cys oxidation. For example, we and others have previously demonstrated increased Cys oxidation in skeletal muscle after fatiguing contractions, suggesting significant physical activity in the day or hours prior to the biopsy could be confounding (Kramer et al., 2018; Pugh et al., 2021). The SOMMA population is predominantly non-Hispanic White, and given our sample size, we have limited ability to test whether associations between signatures of Cys thiol oxidation and our functional outcomes vary by race or ethnicity. Additional factors related to the qualities of muscle, such as fiber type distribution, capillary density, or systemic inflammation can also influence the results. The cross-sectional design allows for testing the concurrent effects of muscle protein oxidation on muscle and physical performance, however repeated assessments are needed to test how Cys oxidation predicts change in performance with aging. Measurements of antioxidant capacity are also warranted, as the levels of antioxidant enzymes (e.g. mitochondrial superoxide dismutase, glutathione peroxidase, catalase) can vary among individuals (Picard, Ritchie, Thomas, Wright, & Hepple, 2011) and may help to interpret our findings. Nonetheless, this first study in older adults has several strengths. It has the power to detect significant associations of muscle protein oxidation with important components of mobility. The state-of-the-art assays included an array of proteins known to play important roles in muscle functions and identified several pronounced signatures of Cys thiol oxidation that associate well with the measures of muscle function, physical performance, and fitness investigated in this study.

In summary, this study supports the hypothesis that higher levels of Cys oxidation on key muscle proteins are largely negatively associated with several measures of muscle function, physical performance, and fitness. These findings should be further tested in a more expanded study or a longitudinal study and investigated in physiological studies to understand the complex impact of cysteine oxidation on muscle and physical function.

## 4. Materials and Methods

### 4.1 Participants and cohort design

The Study of Muscle, Mobility and Aging (SOMMA; https://www.sommaonline.ucsf.edu) is a cohort study of 879 men and women aged 70 or older who were recruited at University of Pittsburgh and Wake Forest University School of Medicine. A description of the design and methods of SOMMA has been published (Cummings et al. 2023). Participants were eligible for this study if they were willing and able to complete a muscle tissue biopsy and magnetic resonance spectroscopy (MRS). All participants provided written and informed consent and the Western IRB-Copernicus Group (WCG) Institutional Review Board approved the SOMMA (WCGIRB #20180764). For this exploratory redox proteomics profiling study, 56 participants were selected partially based on tissue sample availability and to ensure 1) equal distribution across the cohort age range and 2) equal numbers of men and women participants (**Table 1**).

### 4.2 Collection of clinical parameters and phenotype data

SOMMA baseline assessments were completed over the course of several days (Cummings et al., 2023). Age, race, and sex were assessed by questionnaire. Body weight was measured on a balance beam or digital scales. To determine usual walking speed, participants were instructed to walk 10 complete laps around a 40-meter course at their usual pace and without overexerting themselves nor any assistive device other than a straight cane. The total time in seconds to walk 400 meters includes the rest time if the participant stopped walking during the test and was used to calculate walking speed in m/sec. Leg extension strength was assessed by a one repetition max (1RM) on a Keiser Air 420 exercise machine, and peak power was measured across the 40-70% range of leg extension.

Cardiorespiratory fitness was assessed by a cardiopulmonary exercise test (CPET). Participants walked for 5 minutes at a preferred walking speed, and progressive symptom-limited exercise protocol ensued with increases in speed (0.5 mph) and incline (2.5%) in 2-minute increments using a modified Balke protocol or a manual protocol. Cardiorespiratory fitness was determined as VO_2_ peak (mL/min), which was identified as the highest 30-second average of VO_2_ (mL/min) achieved.

### 4.3 Muscle biopsy sample collection

Percutaneous biopsies were collected form the middle region of the musculus vastus lateralis under local anesthesia using a Bergstrom canula with suction (Evans, Phinney, & Young, 1982). Following this, the specimen was blotted dry of blood and interstitial fluid and dissected free of any connective tissue and intermuscular fat. Samples were flash frozen in liquid nitrogen (LN) and stored in the vapor phase of LN. Samples were shipped to PNNL on dry ice for proteomics analysis.

### 4.4 Redox proteomics and mass spectrometry

Muscle biopsy samples (20-30mg) were processed as previously described (Day et al., 2022). 2 additional samples were generated by pooling tissue from the other samples to quantify both oxidized and reduced protein thiols (“total thiol”, **Supplemental Fig. 1A**). After acetone precipitation, the proteins were resuspended in a 200mM Tris (pH 8); 8M Urea buffer, reduced with DTT, and digested overnight with sequence-grade trypsin (Promega) at a 1:50 (µg trypsin: µg protein) ratio. The peptides were cleaned up by C-18 SPE and 100 µg of peptide from each sample was labeled with TMT18plex reagents (ThermoFisher; **Supplemental Fig. 1B**). 133C and 134N were not used for quantification of total oxidation due to potential interference from the 134C and 135N tags used to quantify total thiol. The labeled peptides were then resuspended in 250mM HEPES pH 8, reduced with DTT at a final concentration of 5mM, and a 100µg aliquot of peptide was collected for global proteomics. The remaining peptide was diluted 6-fold with 25mM HEPES pH 7.7 to obtain a DTT concentration of ∼0.8mM to permit enrichment of Cys-containing peptides with thiol affinity resin. The enrichment procedure is adapted from our RAC-TMT workflow (Guo et al., 2014). After enrichment, the enriched redox peptides and global peptides were alkylated with iodoacetamide at 4 times the molarity of the DTT present in the samples and desalted using C-18 SPE.

20µg of enriched redox peptides were collected into 12 concatenated fractions as previously described (Day et al., 2022). Instrument runs were grouped by fraction order within each plex and arranged using a block design strategy (i.e., block 1 = fraction 1 from each plex) to avoid introduction of confounding factors due to decay of instrument performance. Conditions and parameters for LC-MS/MS runs are described elsewhere (Day et al., 2023). Samples of global peptide from each plex were prepared at a concentration of 0.1µg/µL and used for single injections that were run on the same instrument as enriched redox peptides.

### 4.5 MS/MS Data processing and statistical analysis

Raw data were searched against a database containing the Uniprot *Homo sapiens* proteome (release 2023_1 with 20, 407 entries) and common contaminants using MS-GF+ with search parameters specified elsewhere (Day et al., 2022). TMT reporter ion intensities were extracted by MASIC as described in (Day et al., 2022). Data from all fractions of redox multiplexes were aggregated together to the site-centric level and summarized using the R tool “PlexedPiper” (https://github.com/PNNL-Comp-Mass-Spec/PlexedPiper). Additional processing steps are as follows. The redox data was corrected for changes in protein abundance by taking the median intensities of un-normalized global peptide-centric data and scaling them to the average median intensity of the redox data, which was then used to center corresponding channels in the redox data. Data were corrected for multiplex-to-multiplex batch effects as well as biopsy collection site using the “ComBat” function in the R package “sva” (Leek, Johnson, Parker, Jaffe, & Storey, 2012) (https://bioconductor.org/packages/release/bioc/html/sva.html). 75% completeness filtering was set as an additional criterion prior to median centering of Cys site expression and the data was filtered for Cys sites corresponding to a select group of proteins related to muscle protein structure and contraction (**Supplemental File S1**) for prioritized analysis.

Statistical analysis was performed using linear regression modeling with the “limma” R package for its enhanced statistical power with low sample size (Smyth, 2005) (https://bioconductor.org/packages/release/bioc/html/limma.html). The models were not stratified by sex due to the limited number of samples and were adjusted for sex, age, and weight of the participants as covariates to test against the phenotypes of leg power, leg strength, walking speed, and cardiopulmonary fitness. Cys sites with statistically significant associations were defined by a p value < 0.05 threshold. For each phenotype, significantly associated Cys sites were plotted on heatmaps if they had 100% completeness across all samples and Cys oxidation levels for patients without phenotype data were excluded from heatmaps. Heatmap phenotypes were converted to Z-score and samples correspondingly arranged prior to row clustering. Heatmaps were segregated into 6 distinct clusters defined by the hierarchical clustering dendrogram. Cluster centroids were defined by averaging feature intensities for each sample within a cluster. Cluster centroids were then plotted against non-Z-scored phenotype to produce cluster correlation plots scored by Pearson correlation. Cluster-to-phenotype correlation was further evaluated by limma using cluster centroids as a combined feature (**Supplemental File S3**).

### 4.6 Data availability

The mass spectrometry data have been deposited to the MassIVE repository with the dataset accession: MSV000093136. SOMMA data are also available at https://sommaonline.ucsf.edu/.

## Data Availability

https://massive.ucsd.edu/ProteoSAFe/private-dataset.jsp?task=df7827acfab9489980eeb4508e7da208

## Acknowledgements

Mass spectrometry proteomics experiments were performed in the Environmental Molecular Sciences Laboratory, Pacific Northwest National Laboratory, a national scientific user facility sponsored by the DOE under Contract DE-AC05-76RL0 1830.

---

**Supplemental Figure S1.**
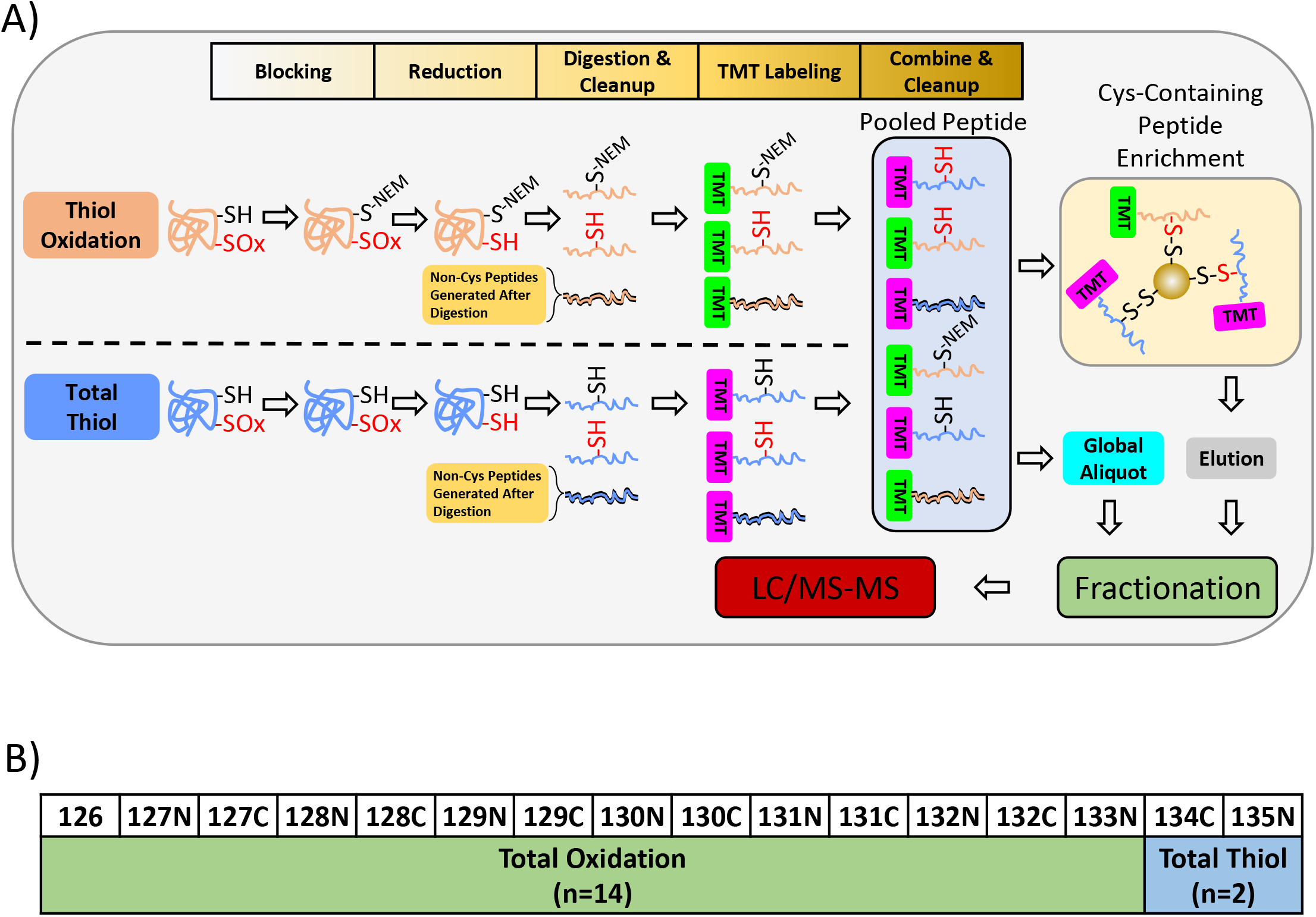
Integrated global and redox proteomics workflow. A) The RAC-TMT workflow (Guo et al., 2014) was modified to integrate both global and redox proteomics workflows into the same processing procedure from the same sample. The blocking and reduction strategies critical for measuring total oxidation of Cys sites were incorporated into a standard global proteomic processing workflow, ultimately allowing for both enrichment of oxidatively modified peptides and global proteomics from the same pool of peptide. The global proteomic data is used to calibrate the redox proteomics data in a similar manner as with phosphoproteomics data (Wu et al., 2011), to distinguish changes in Cys oxidation from changes in protein expression. Off-line reverse phase high pressure liquid chromatography fractionation of enriched redox peptides was implemented to increase coverage, as was recently demonstrated in one of our previous reports (Day et al., 2022). Total thiol channels were incorporated for a signal boosting strategy as described in (Day et al., 2022), as well as to provide enough peptide material for offline fractionation. B) Representative TMT18plex design. Muscle biopsy samples were randomly assigned different channels within a multiplex. A total of 4 plexes were generated to analyze 56 samples, where pooled samples were used to generate total thiol samples within each plex.

**Supplemental Figure S2.**
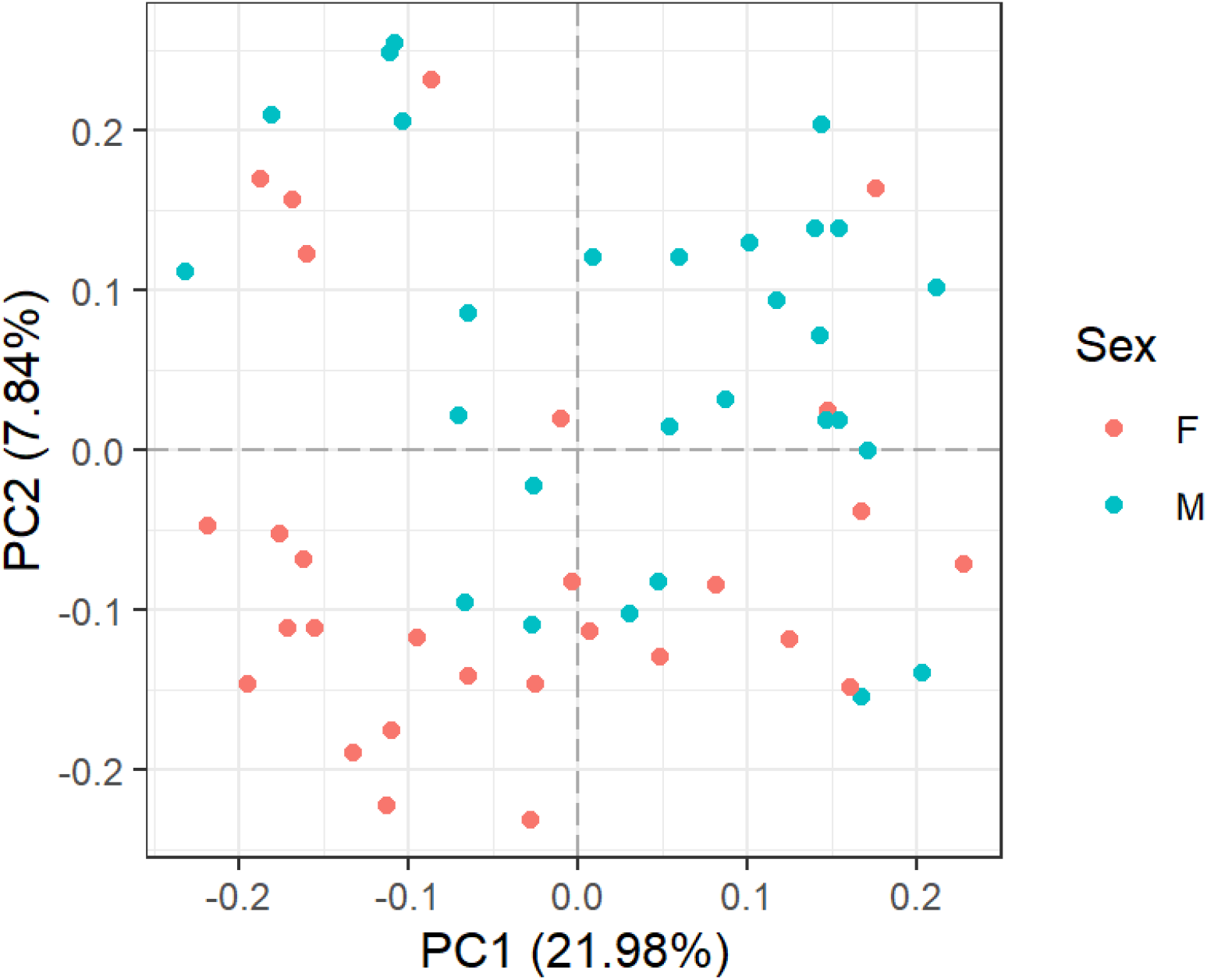
Variation in muscle protein oxidation among participants. Representative principal component analysis (PCA) plot of redox proteomics data with each data point (participant) colored by sex.

**Supplemental Figure S3.**
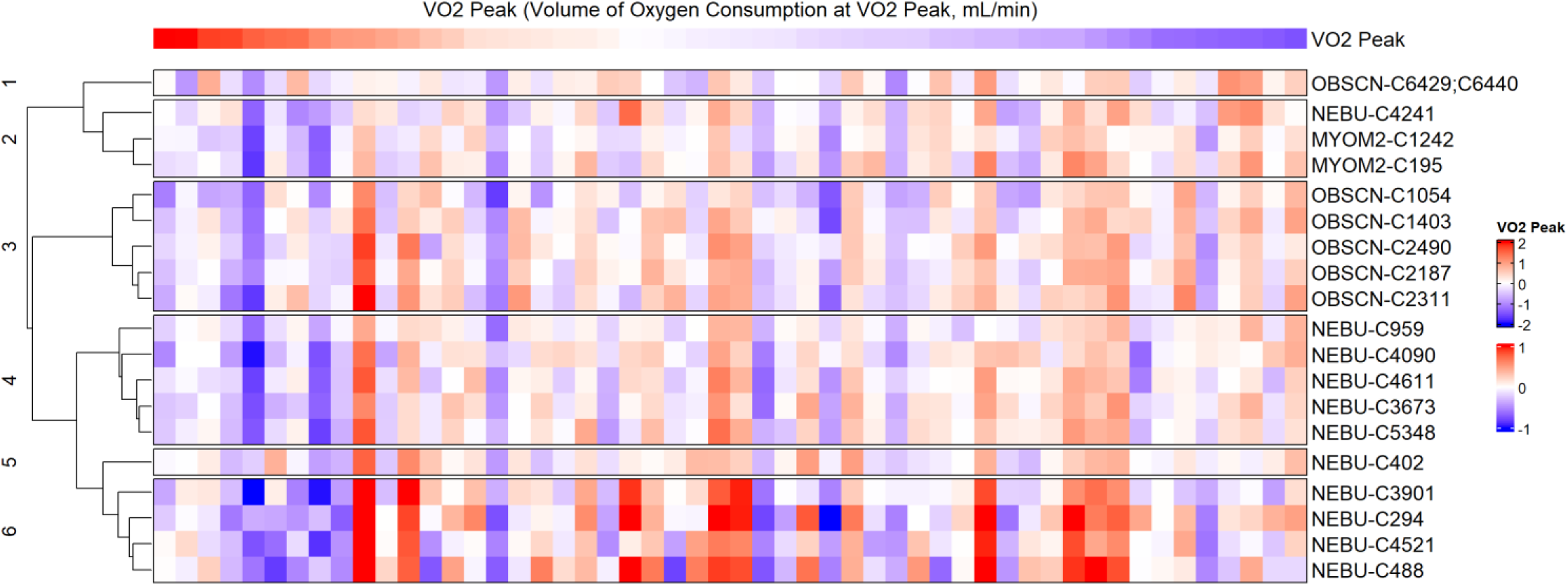
Heatmaps and correlation plots of all Cys sites that are significantly associated with the four phenotypes investigated. For the heatmaps (**Supplemental Figures S3, S5, S7, S9**), each row represents a Cys site with oxidation levels that are significantly associated with a phenotype, while each column represents a participant. Columns are ranked in descending order from left to right based on phenotypic measurement and the phenotypic values are represented by median-centered Z-scores, while Cys oxidation levels were scaled by median-centering. Protein identities are in UniProt format. Sites passing an adjusted p value < 0.05 cutoff are denoted by ‘*’. For each signature (sub-cluster) in a heatmap, Cys sites were plotted for correlation analysis (**Supplemental Figures S4, S6, S8, S10**), where the mean level of oxidation across all Cys sites in a signature were plotted for each participant against their phenotypic measurement. *R* and p values are derived from Pearson correlation.

**Supplemental Figure S4.**
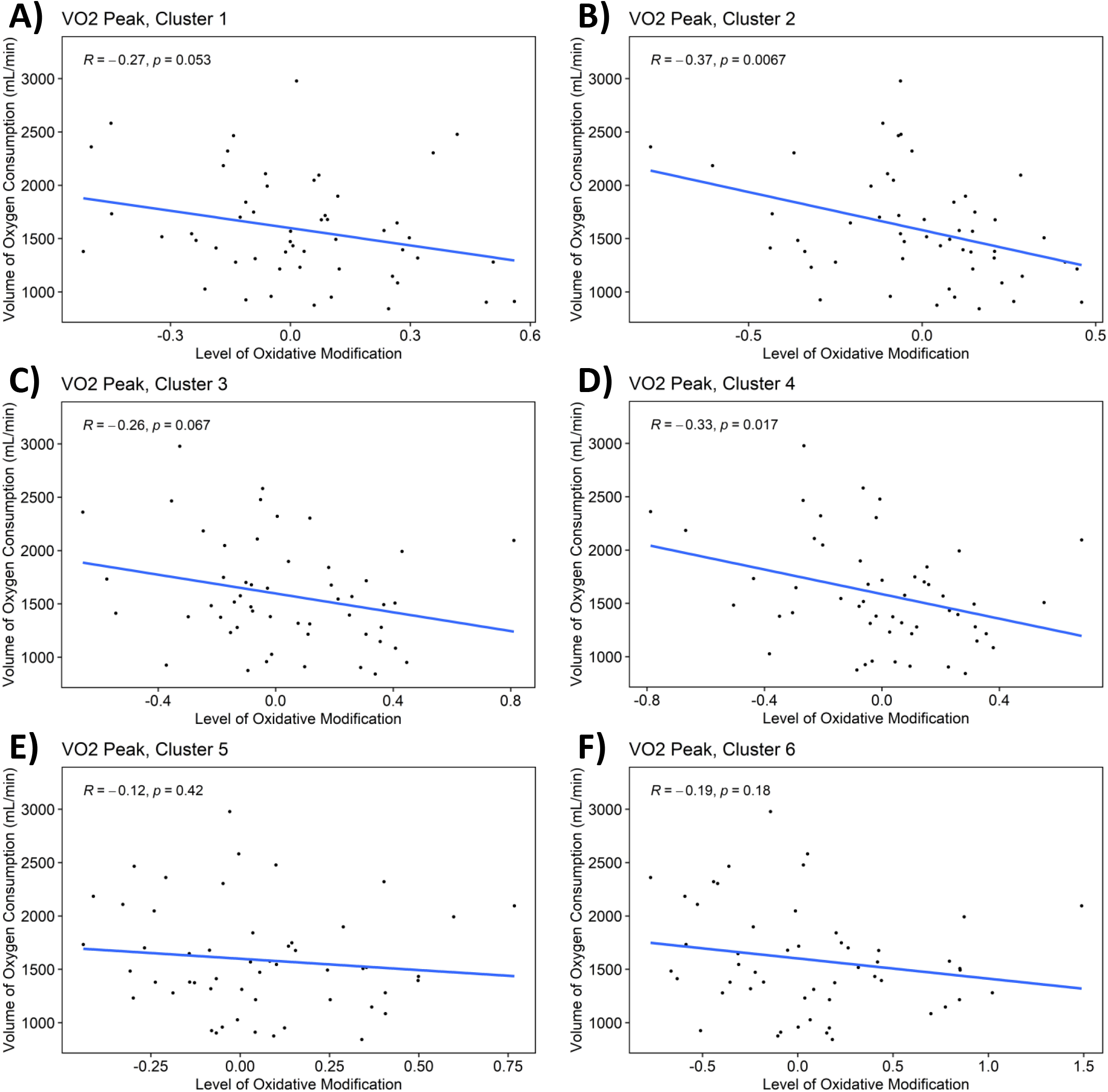
Heatmaps and correlation plots of all Cys sites that are significantly associated with the four phenotypes investigated. For the heatmaps (**Supplemental Figures S3, S5, S7, S9**), each row represents a Cys site with oxidation levels that are significantly associated with a phenotype, while each column represents a participant. Columns are ranked in descending order from left to right based on phenotypic measurement and the phenotypic values are represented by median-centered Z-scores, while Cys oxidation levels were scaled by median-centering. Protein identities are in UniProt format. Sites passing an adjusted p value < 0.05 cutoff are denoted by ‘*’. For each signature (sub-cluster) in a heatmap, Cys sites were plotted for correlation analysis (**Supplemental Figures S4, S6, S8, S10**), where the mean level of oxidation across all Cys sites in a signature were plotted for each participant against their phenotypic measurement. *R* and p values are derived from Pearson correlation.

**Supplemental Figure S5.**
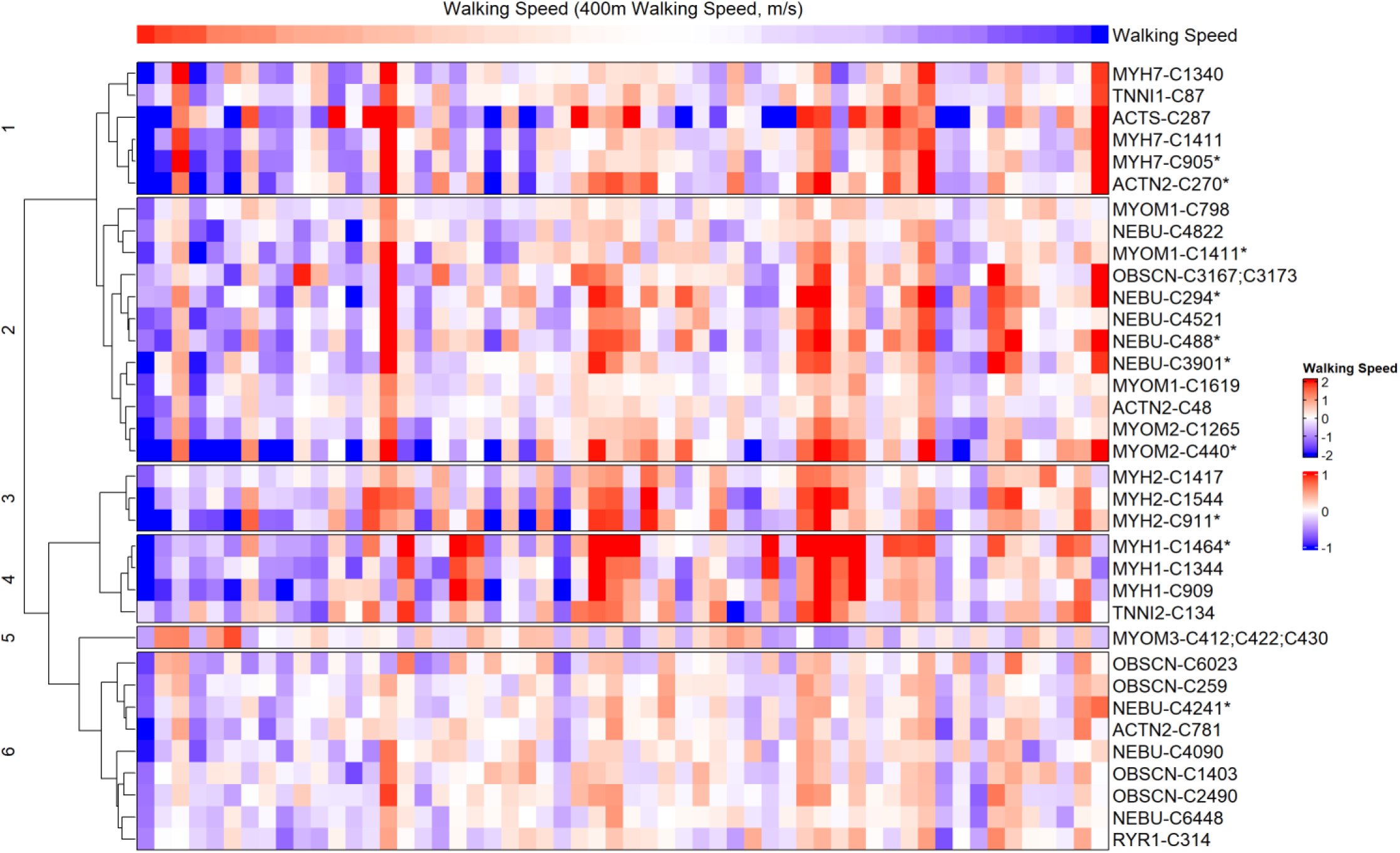
Heatmaps and correlation plots of all Cys sites that are significantly associated with the four phenotypes investigated. For the heatmaps (**Supplemental Figures S3, S5, S7, S9**), each row represents a Cys site with oxidation levels that are significantly associated with a phenotype, while each column represents a participant. Columns are ranked in descending order from left to right based on phenotypic measurement and the phenotypic values are represented by median-centered Z-scores, while Cys oxidation levels were scaled by median-centering. Protein identities are in UniProt format. Sites passing an adjusted p value < 0.05 cutoff are denoted by ‘*’. For each signature (sub-cluster) in a heatmap, Cys sites were plotted for correlation analysis (**Supplemental Figures S4, S6, S8, S10**), where the mean level of oxidation across all Cys sites in a signature were plotted for each participant against their phenotypic measurement. *R* and p values are derived from Pearson correlation.

**Supplemental Figure S6.**
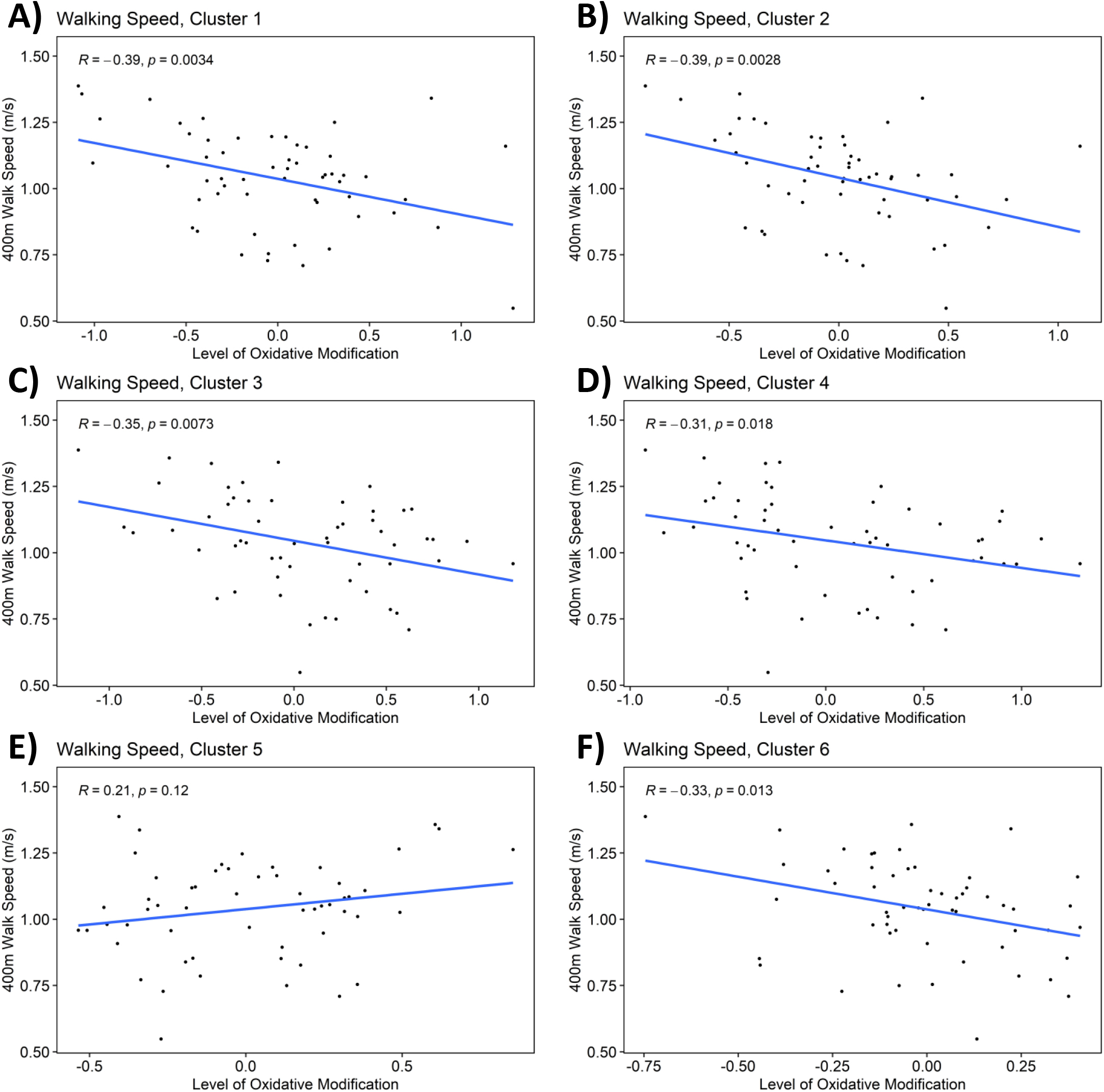
Heatmaps and correlation plots of all Cys sites that are significantly associated with the four phenotypes investigated. For the heatmaps (**Supplemental Figures S3, S5, S7, S9**), each row represents a Cys site with oxidation levels that are significantly associated with a phenotype, while each column represents a participant. Columns are ranked in descending order from left to right based on phenotypic measurement and the phenotypic values are represented by median-centered Z-scores, while Cys oxidation levels were scaled by median-centering. Protein identities are in UniProt format. Sites passing an adjusted p value < 0.05 cutoff are denoted by ‘*’. For each signature (sub-cluster) in a heatmap, Cys sites were plotted for correlation analysis (**Supplemental Figures S4, S6, S8, S10**), where the mean level of oxidation across all Cys sites in a signature were plotted for each participant against their phenotypic measurement. *R* and p values are derived from Pearson correlation.

**Supplemental Figure S7.**
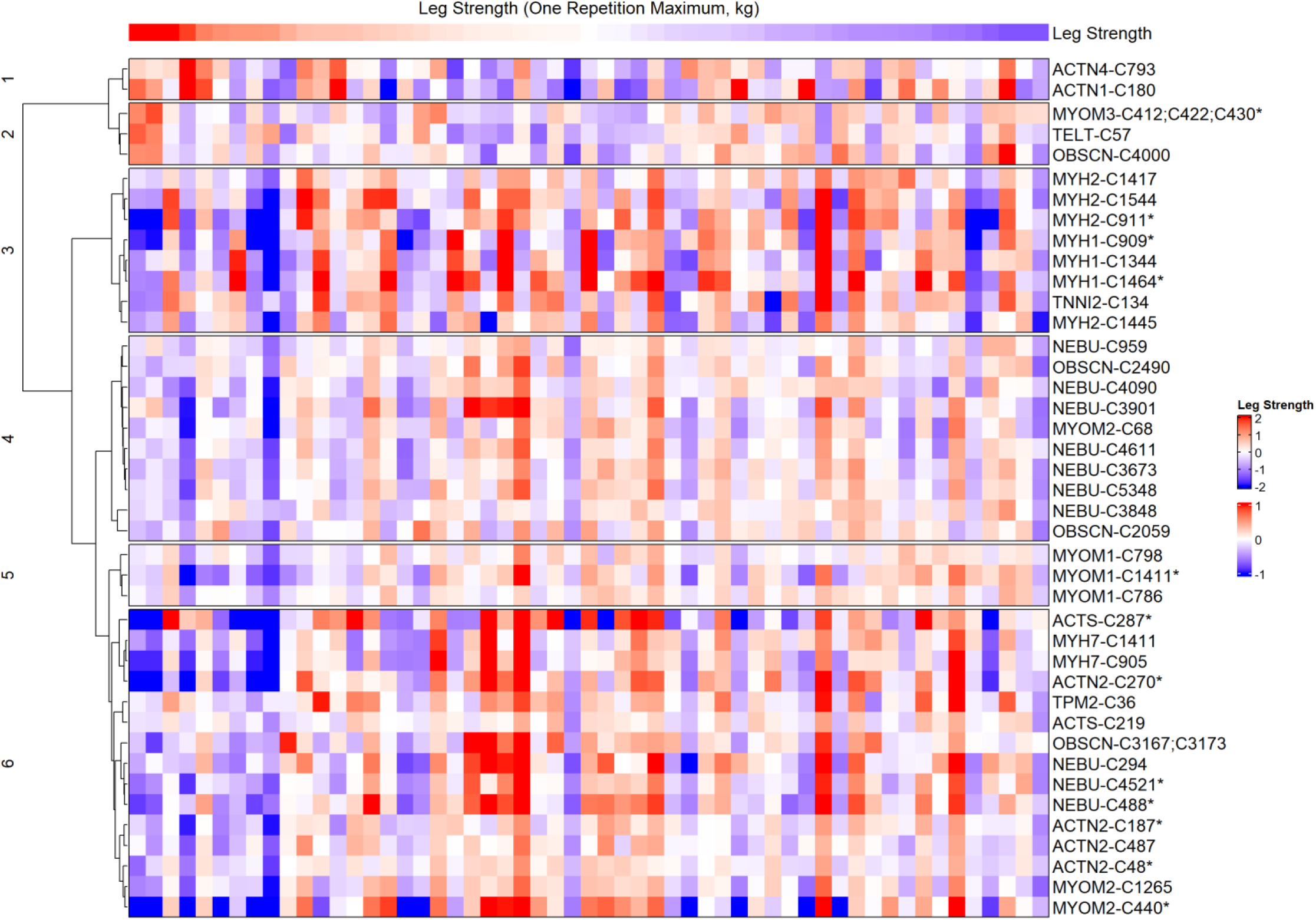
Heatmaps and correlation plots of all Cys sites that are significantly associated with the four phenotypes investigated. For the heatmaps (**Supplemental Figures S3, S5, S7, S9**), each row represents a Cys site with oxidation levels that are significantly associated with a phenotype, while each column represents a participant. Columns are ranked in descending order from left to right based on phenotypic measurement and the phenotypic values are represented by median-centered Z-scores, while Cys oxidation levels were scaled by median-centering. Protein identities are in UniProt format. Sites passing an adjusted p value < 0.05 cutoff are denoted by ‘*’. For each signature (sub-cluster) in a heatmap, Cys sites were plotted for correlation analysis (**Supplemental Figures S4, S6, S8, S10**), where the mean level of oxidation across all Cys sites in a signature were plotted for each participant against their phenotypic measurement. *R* and p values are derived from Pearson correlation.

**Supplemental Figure S8.**
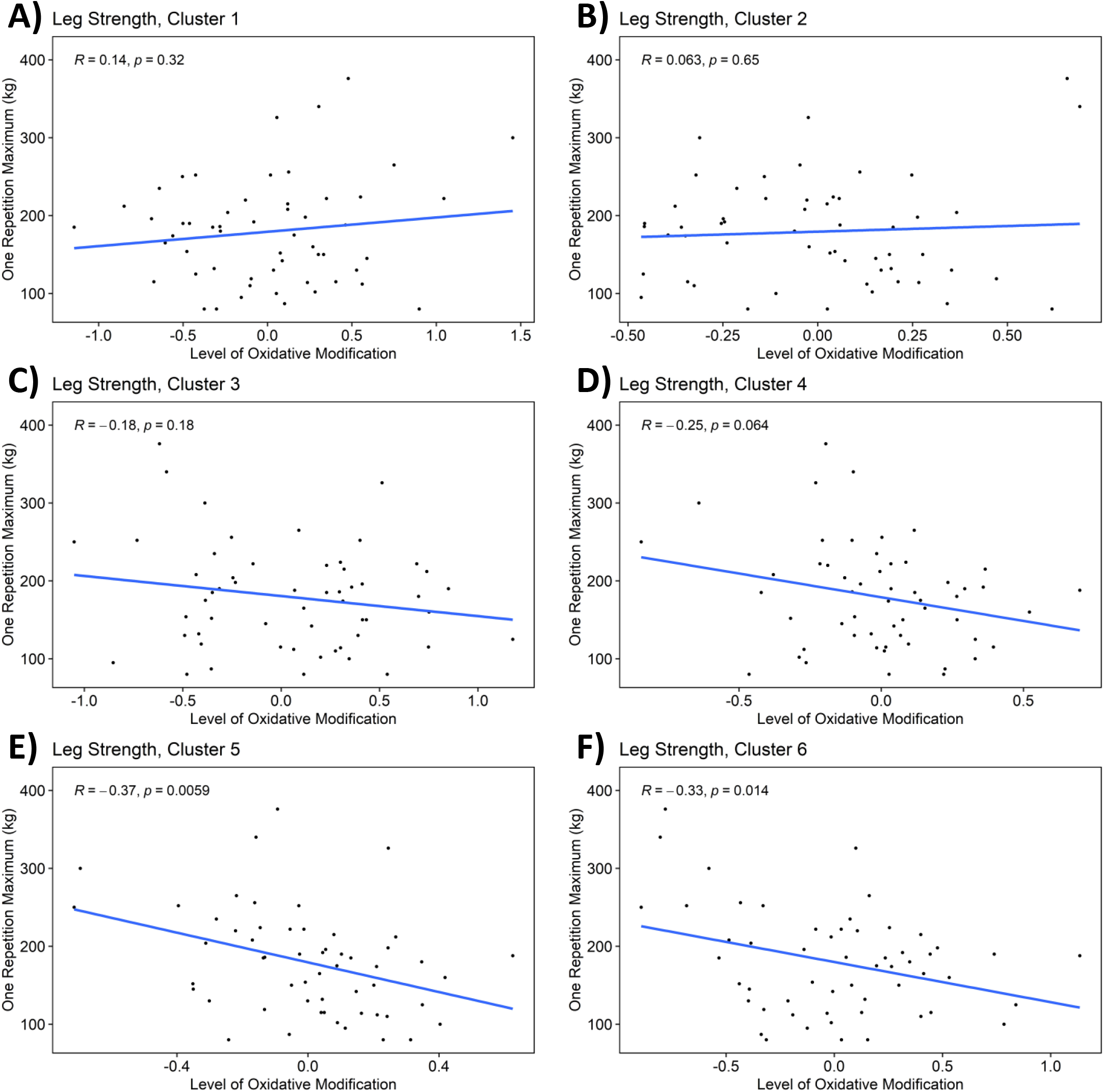
Heatmaps and correlation plots of all Cys sites that are significantly associated with the four phenotypes investigated. For the heatmaps (**Supplemental Figures S3, S5, S7, S9**), each row represents a Cys site with oxidation levels that are significantly associated with a phenotype, while each column represents a participant. Columns are ranked in descending order from left to right based on phenotypic measurement and the phenotypic values are represented by median-centered Z-scores, while Cys oxidation levels were scaled by median-centering. Protein identities are in UniProt format. Sites passing an adjusted p value < 0.05 cutoff are denoted by ‘*’. For each signature (sub-cluster) in a heatmap, Cys sites were plotted for correlation analysis (**Supplemental Figures S4, S6, S8, S10**), where the mean level of oxidation across all Cys sites in a signature were plotted for each participant against their phenotypic measurement. *R* and p values are derived from Pearson correlation.

**Supplemental Figure S9.**
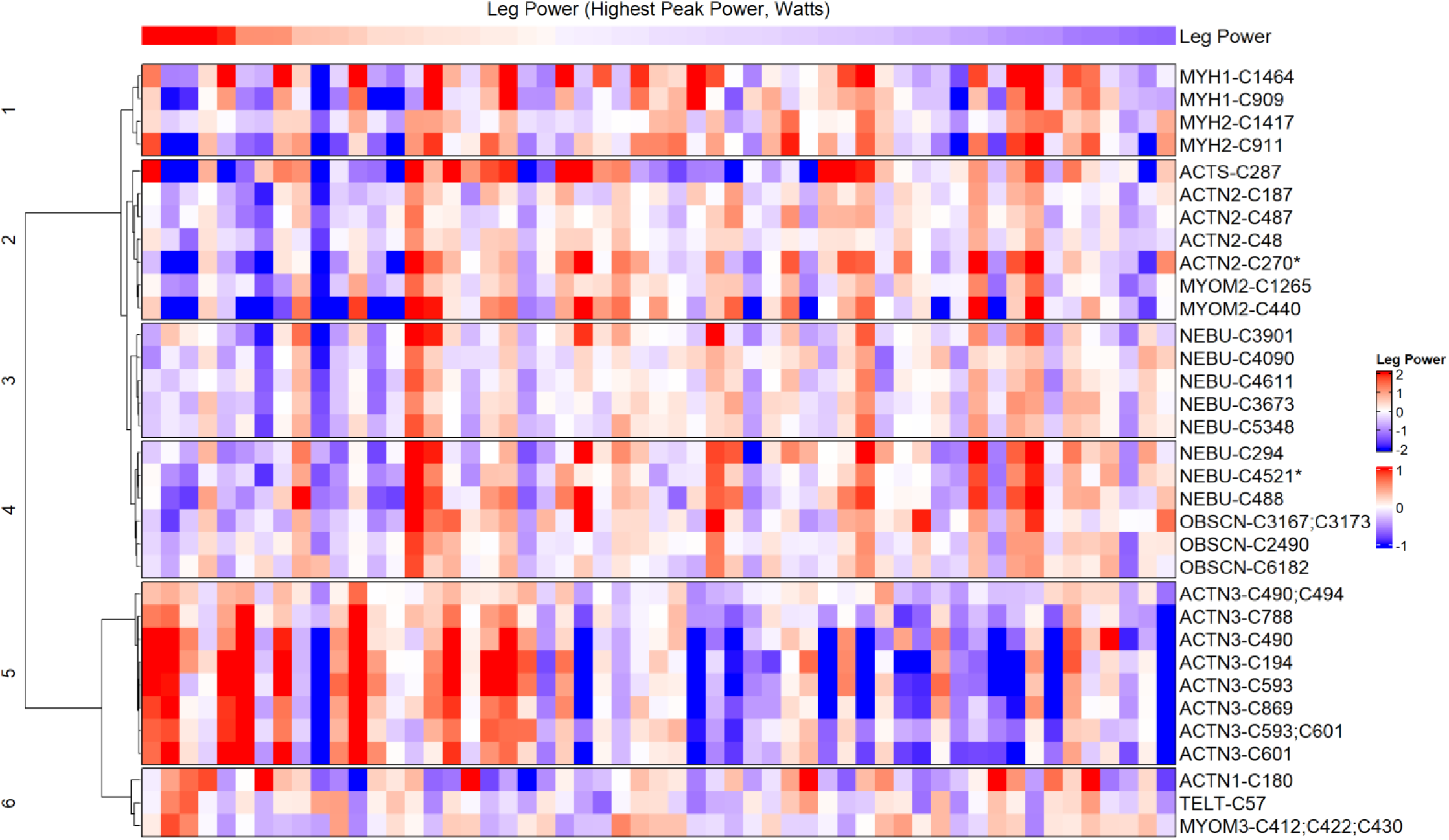
Heatmaps and correlation plots of all Cys sites that are significantly associated with the four phenotypes investigated. For the heatmaps (**Supplemental Figures S3, S5, S7, S9**), each row represents a Cys site with oxidation levels that are significantly associated with a phenotype, while each column represents a participant. Columns are ranked in descending order from left to right based on phenotypic measurement and the phenotypic values are represented by median-centered Z-scores, while Cys oxidation levels were scaled by median-centering. Protein identities are in UniProt format. Sites passing an adjusted p value < 0.05 cutoff are denoted by ‘*’. For each signature (sub-cluster) in a heatmap, Cys sites were plotted for correlation analysis (**Supplemental Figures S4, S6, S8, S10**), where the mean level of oxidation across all Cys sites in a signature were plotted for each participant against their phenotypic measurement. *R* and p values are derived from Pearson correlation.

**Supplemental Figure S9.**
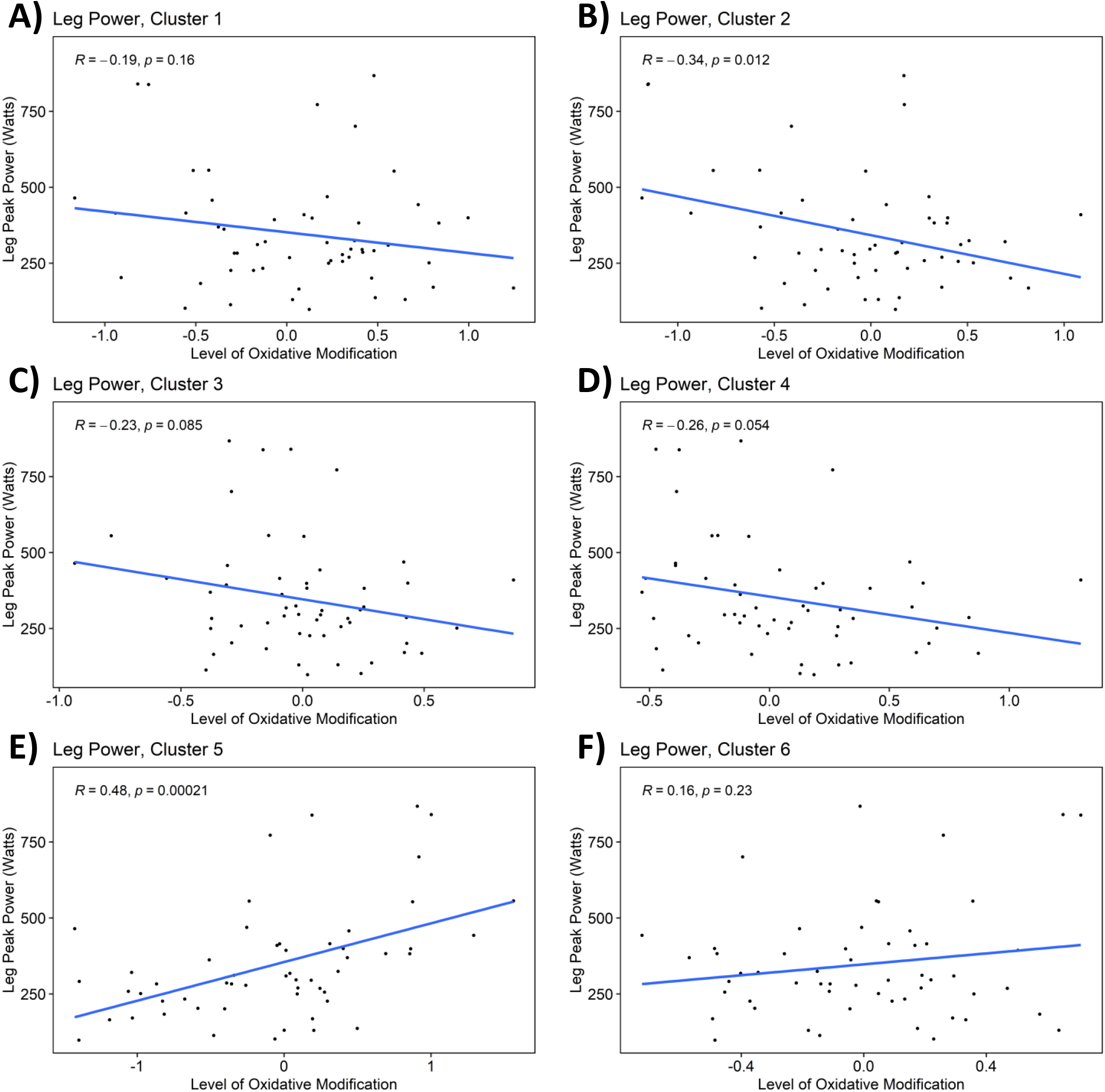
Heatmaps and correlation plots of all Cys sites that are significantly associated with the four phenotypes investigated. For the heatmaps (**Supplemental Figures S3, S5, S7, S9**), each row represents a Cys site with oxidation levels that are significantly associated with a phenotype, while each column represents a participant. Columns are ranked in descending order from left to right based on phenotypic measurement and the phenotypic values are represented by median-centered Z-scores, while Cys oxidation levels were scaled by median-centering. Protein identities are in UniProt format. Sites passing an adjusted p value < 0.05 cutoff are denoted by ‘*’. For each signature (sub-cluster) in a heatmap, Cys sites were plotted for correlation analysis (**Supplemental Figures S4, S6, S8, S10**), where the mean level of oxidation across all Cys sites in a signature were plotted for each participant against their phenotypic measurement. *R* and p values are derived from Pearson correlation.

**Supplemental File S1**. Muscle sarcomere structure and contraction proteins investigated in this study.

**Supplemental File S2**. Results of linear regression analyses (limma) testing associations between protein Cys thiol oxidation sites and phenotypic measurements.

**Supplemental File S3**. Results of linear regression analyses (limma) testing associations between protein Cys thiol oxidation signatures and phenotypic measurements.

